# Assessment and clinical utility of a non-Next-Generation Sequencing based Non-Invasive Prenatal Testing technology

**DOI:** 10.1101/2021.06.21.21256776

**Authors:** U Gormus, A Chaubey, S Shenoy, YW Wong, LY Chan, BP Choo, L Oraha, A Gousseva, F Persson, L Prensky, E Chin, M Hegde

## Abstract

**Background:** Rolling circle replication (RCR) is a novel technology that has not been applied to cell-free DNA (cfDNA) testing until recently. Given the cost and simplicity advantages of this technology compared to other platforms currently used in cfDNA analysis, an assessment of RCR in clinical laboratories was performed. Here, we present the first validation study from clinical laboratories utilizing RCR technology.

**Methods:** 831 samples from spontaneously pregnant women carrying a singleton fetus and 25 synthetic samples were analyzed for the fetal risk of Trisomy 21, Trisomy 18 and Trisomy 13 by three laboratories on three continents. All women who provided the samples were followed to birth, where evaluation for fetal aneuploidies was performed using newborn examinations and any suspected aneuploidies were confirmed with karyotyping.

**Results:** The study found rolling circle replication to be a highly viable technology for clinical assessment of fetal aneuploidies with 100% sensitivity for T21 (95% CI:82.35% - 100.00%); 100.00% sensitivity for T18 (71.51% - 100.00%) and 100.00% sensitivity for T13 analyses (66.37% - 100.00%). The specificities were >99% for each trisomies [99.7% (99.01% - 99.97%) for T21; 99.5% (98.62% - 99.85%) for T18; 99.7% (99.03% - 99.97%) for T13], along with a first pass no-call rate of 0.93%.

**Conclusions:** The study showed that using a rolling circle replication-based cfDNA system for the evaluation of the common aneuploidies would provide greater accuracy and clinical utility compared to conventional biochemical screening and comparable results to other reported cfDNA methodologies.

## INTRODUCTION

Trisomies are important chromosomal aberrations often associated with varying degrees of intellectual disabilities, several health and developmental defects, and whose incidence is correlated with increasing maternal age[1]. Although the average maternal age has increased globally in the last 50 years, the incidence of trisomy has significantly decreased during that time frame due to the increased utilization of improved prenatal screening tests[2]. Historically, these prenatal screening tests consisted of biochemical blood tests and/or ultrasound scans. These conventional screening tests are still used globally, but due to their higher false positive rates and lower detection rates, they have started to be replaced by newer, more accurate technologies using the placental cell-free DNA (cfDNA) circulating in the maternal blood. Cell-free nucleic acids, also known as extracellular nucleic acids, are fragments of DNA or RNA molecules that are released from cells into the body fluids.

Lo et al were the first to report that a portion of the cell-free DNA in maternal blood was from the fetus and placenta and to comment on how cell-free fetal DNA was suitable for prenatal examinations[3]. The introduction of cell-free DNA into prenatal clinical practice first started through the use of next-generation sequencing (NGS) technology for the assessment of Trisomy 21 (T21), Trisomy 18 (T18) and Trisomy 13 (T13), and was referred to as Non-Invasive Prenatal Testing (NIPT)[4,5]. Although NIPT has been shown to be highly accurate, the next generation sequencing techniques that were used has limited the global accessibility to this test due to its high cost and complexity. It has been noted that a considerable cost reduction is necessary to make this approach cost effective enough to be commonly used[6]. Furthermore, the complexity of the NGS-based technologies adds additional hurdles to the ability of laboratories to implement this test. Vanadis^®^ NIPT was developed without using NGS or polymerase chain reaction (PCR), to enable a cost effective and high performance cfDNA aneuploidy screening.

Vanadis^®^ NIPT is a new technology targeting relevant chromosomes based on a digital molecular quantification in a 96□well microplate[7,8]. The method converts targeted chromosomal fragments into digitally quantifiable objects through rolling circle replication and chromosome-specific labeling. The normalized ratio between the number of chromosome specific objects are then used to calculate the z-score which is mapped to a post-test risk.

Here, we report on the clinical performance of the Vanadis^®^ NIPT assay in PerkinElmer Genomics Laboratories.

## MATERIALS AND METHOD

### Ethics Statement

Protocols used for sample collection were approved by the Research Ethics Board of CHU de Québec (#2016-2989 and #2020-4895). The study was performed in accordance with the ethical standards of the institutional and/or national research committees.

### Study Population and Clinical Evaluation

Validation protocols were written based upon templates relevant to the Vanadis system (Supplemental A and B). Based on this, a total aggregated set of 831 samples from spontaneously pregnant women carrying a singleton fetus were analyzed. The inclusion criteria for participation in this study were pregnant women between the ages of 18 and 50 and between 10 and 40 gestational weeks. The women were not selected based on prior risk and all consented to participate in the study. All subjects were followed to birth, where evaluation for fetal aneuploidies was performed using newborn examinations and any suspected aneuploidies were confirmed with karyotyping. 10 milliliters (mL) of blood were collected from each woman between February 2019 and July 2019 at maternity clinics in Kuala Lumpur and Quebec. Blood samples were processed as described below, and at least 3 mL of plasma was extracted and sent to PerkinElmer Genomics (PKIG) labs located in Sollentuna, Sweden; Kuala Lumpur, Malaysia; and Pittsburgh, USA.

Ten samples from confirmed T21 positive pregnancies and three samples from confirmed T18 positive pregnancies and one sample from a confirmed T13 pregnancy were used. Other trisomy positive control samples (nine of T21, eight of T18 and eight of T13) were purchased from SeraCare Life Sciences, Inc. (USA) (Seraseq Trisomy 21 aneuploidy reference material-0720-0019, Trisomy 13 aneuploidy reference material-0720-0017, Trisomy 18 aneuploidy reference material-0720-0018).

### Sample collection and preparation

Blood samples were collected into Cell-FreeTM DNA BCT tubes (Streck, Omaha, USA) from each pregnant woman. After arrival in the lab by courier, study samples were barcoded with unique subject codes and patient identification numbers and anonymized.

Samples were processed in the PKIG lab in Kuala Lumpur and the CHU de Québec-Université Laval lab in Quebec by using a double centrifugation protocol[8]. All plasma was separated within 5 days of blood draw and stored in new plasma storage tubes. The plasma tubes were barcoded with unique subject codes and patient identification numbers were anonymized. The plasma tubes were stored at −80°C until processing at a PKIG Laboratory.

### Test method

Samples were analyzed using the Vanadis^®^ system following existing manuals and instructions for use. The Vanadis^®^ NIPT assay uses a series of enzymatic steps to generate labelled rolling circle replication products (RCPs) from chromosomal cfDNA targets, as previously described[7]. Automated extraction of cfDNA from plasma was performed using the Vanadis Extract^®^ platform, followed by continued processing on the Vanadis Core^®^ platform to generate labelled RCPs, which were then imaged and counted using the Vanadis View^®^ instrument. The performance metrics to be evaluated were based on the Z-score results were calculated with Lifecycle^™^ software version 7.2 and exported to an Excel file.

### Data Analysis and Sample Classification

Automated data analysis and quality assessment were performed, and chromosomal ratio calculations were calculated for all approved samples. The results were classified into low or high risk with a Z□score approach based on each normalized chromosomal ratio and the sample[specific standard deviation. The Z-score cut-off values were 3.5 for chromosome 21 and 3.15 for chromosomes 18 and 13. The samples that failed the quality assessment were rejected and classified as ‘no□call’. The fetal sex was classified from the number of detected RCPs from chromosome Y relative to the number of RCPs from the measured autosomal chromosomes using an adaptive binary classifier[7,8]. Measured fetal fraction, which is often thought to be a useful quality control metric, was not gathered as recent studies have shown that it can be significantly incorrect[9].

## RESULTS

A total of 856 samples (Figure 1) were included in the study, 831 of them were taken from singleton pregnancies with spontaneous fertilization and 25 were reference material provided by SeraCare Life Sciences, Inc. (USA). There were eight first pass no call results that were excluded from calculations (first pass no call rate: 0.93%). The average median maternal age in the study group was 32 (min:20 years, max: 46 years). The median gestational age was 12 weeks 5 days (min:10 weeks, max: 34 weeks).

**Figure 1.**
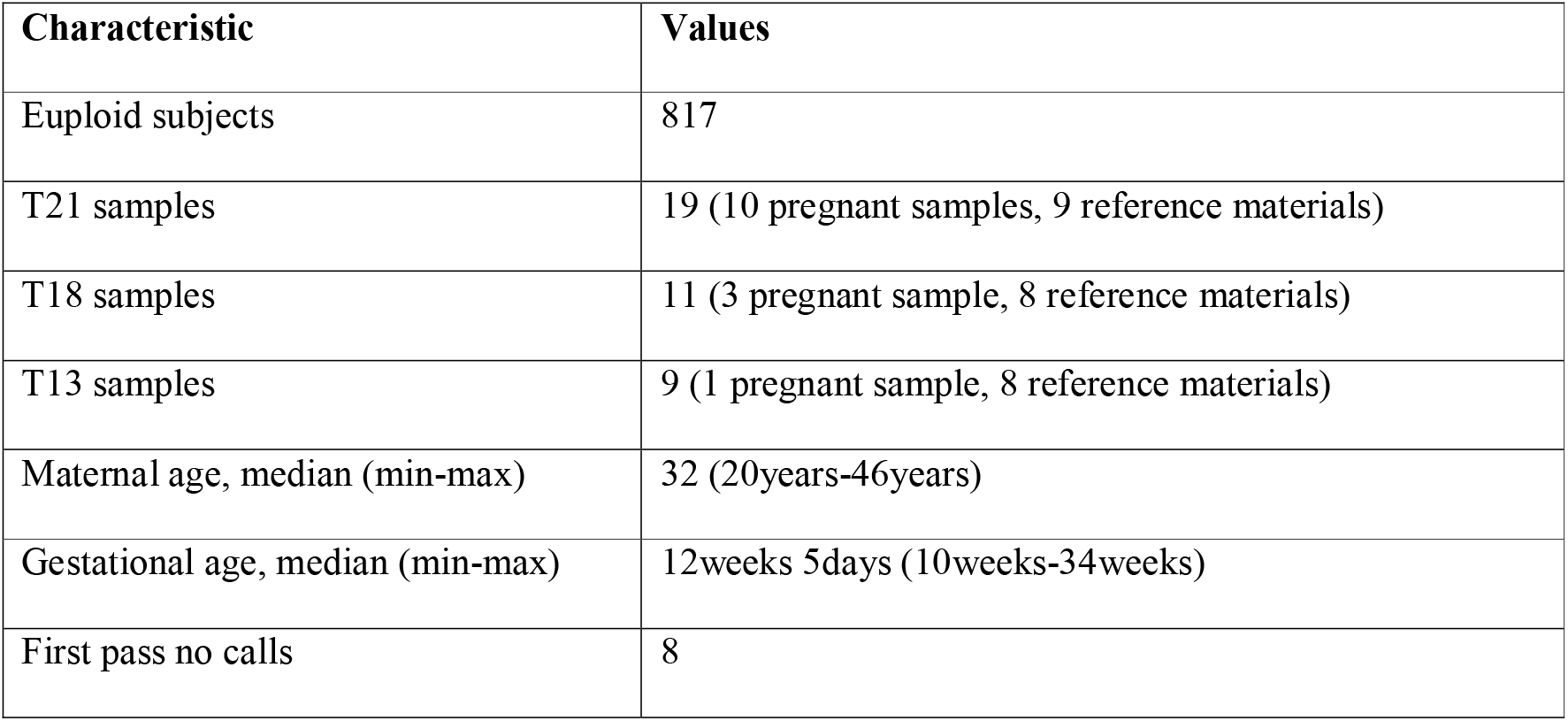
Characteristics of Study Subjects:

The results from the test (Table 1) showed 100% sensitivity for T21 [95% Confidence Interval (CI):82.35% - 100.00%]; 100.00% sensitivity for T18 (95% CI:71.51% - 100.00%) and 100.00% sensitivity for T13 analyses (95% CI:66.37% - 100.00%). The specificities were >99% for each trisomies [99.7% (95% CI: 99.01% - 99.97%) for T21; 99.5% (95% CI:98.62% - 99.85%) for T18; 99.7% (95% CI:99.03% - 99.97%) for T13].

**Table one:** Test performance Vanadis^®^ NIPT – Aneuploidy (Sweden+Malaysia+USA)

|  | **Trisomy 21** | **Trisomy 18** | **Trisomy 13** |
| --- | --- | --- | --- |
| **Total subjects** | 408+214+234=856 | 408+214+234=856 | 408+214+234=856 |
| **No calls:** | 8 (no call rate: 0.93%, with unrepeated samples) |  |  |
| **Without no calls:** | 848 | 848 | 848 |
| **True positives †** | 7(5)+4(0)+8(4)=19 | 7(6)+2(0)+2(2)=11 | 6(6)+1(0)+2(2)=9 |
| **False positives** | 2+0+0=2 | 4+0+0=4 | 1+0+1=2 |
| **True negatives** | 827 | 833 | 837 |
| **False negatives** | 0 | 0 | 0 |
| **Sensitivity (95% CI)** | 100.00% (82.35% to 100.00%) | 100.00% (71.51% to 100.00%) | 100.00% (66.37% to 100.00%) |
| **Specificity (95% CI)** | 99.76% (99.13% to 99.97%) | 99.52% (98.78% to 99.87%) | 99.76% (99.14% to 99.97%) |

†25 out of 39 are SeraCare samples; SeraCare samples are within parentheses

No false negative results were detected (FNR: 0%) with low levels of false positive rates (FPR: 0.24% for T21, 0.47% for T18 and 0.24% for T13).

For fetal gender assessment, accuracy was 98.80% (Table 2). Of note, a recent blinded study at an independent site using improved Y chromosome detection modifications in reagents and analysis software showed 100% concordance for fetal sex determination between Vanadis and NGS methods (n= 251 samples; unpublished observations).

**Table two:** Test performance Vanadis^®^ NIPT – Sex classification

| Sweden |  |  | Malaysia |  |  | USA |  |  |
| --- | --- | --- | --- | --- | --- | --- | --- | --- |
| 391 |  |  | 214 |  |  | 234 |  |  |
|  | Females | Males |  | Females | Males |  | Females | Males |
| Total subjects | 166 | 225 | Total subjects | 94 | 120 | Total subjects | 101 | 133 |
| No calls | 6 |  | No calls | 2 |  | No calls | 0 |  |
| Total subjects (w/o no calls) | 164 | 221 | Total subjects (w/o no calls) | 92 | 120 | Total subjects (w/o no calls) | 101 | 133 |
| Correct classification | 162 | 220 | Correct classification | 92 | 120 | Correct classification | 98 | 129 |
| Incorrect classification | 2 | 1 | Incorrect classification | 0 | 0 | Incorrect classification | 3 | 4 |
|  | Females | Males | TOTAL | Performance Criteria | Females | Males | TOTAL |  |
| Total subjects | 361 | 478 | 839 | Accuracy | 98.79% | 98.79% | 98.79% |  |
| No calls excluded: | 357 | 474 | 831 |  |  |  |  |  |
| Correct classification | 352 | 469 | 821 |  |  |  |  |  |
| Incorrect classification | 5 | 5 | 10 |  |  |  |  |  |

## DISCUSSION

Vanadis^®^ NIPT is an efficient and cost-effective option for prenatal screening. The test can be offered to pregnant women starting from the 10th week of gestation and can be integrated as a first tier choice as prenatal screening analysis as it is more cost-effective than the NGS-based NIPT[10] and has a higher sensitivity and specificity compared to the conventional biochemical screening[11].

This study shows high sensitivity and specificity of Vanadis NIPT analysis. In this sample set, all aneuploidy cases were detected accurately, thus resulting in a sensitivity of 100% for trisomy 21, trisomy 18, and trisomy 13 and a ≥99.5% specificity. Specificity would likely be even higher if a second tube of blood was available for the samples with borderline Z-scores. Furthermore, if a second sample was available for these patients, then the low first pass no call rate of 0.93% would likely be reduced to a final no call rate of around 0.1%, based upon a previous study showing a 87.5% reduction of no calls when a second sample is run on the Vanadis^®^ system[12].

Studies have shown that the sensitivity and specificity of NIPT are better than the conventional screening methods [13-21] which has lead professional societies (such as the American College of Obstetricians and Gynecologists and the Society for Maternal-Fetal Medicine) to state “Cell-free DNA is the most sensitive and specific screening test for the common fetal aneuploidies”[11]. NIPT technologies that involve next generation sequencing have shown that 98 - 100% of common aneuploidies can be detected at a combined false positive rate of 0.44 - 0.91% [22]), while conventional biochemical screening can range from 50 - 95%, with a false positive rate of 5%, depending upon which screening strategy was used[23]. By providing higher detection rates and lower false positive and negative rates compared to conventional screening, NIPT technologies are more clinically effective and lead to fewer invasive procedures[24].

As this study shows, the Vanadis^®^ system provided results comparable to those of the more common NIPT technologies (Table 3). Both groups show similar sensitivities and specificities, which are greater than those for the conventional biochemical screening, thus emphasizing their clinical utility. Although similar in performance, there is a difference when it comes to the technological complexity and cost-effectiveness. By removing the need for PCR and NGS, the installation, hands-on time, bioinformatics and run costs are automatically significantly lower with the Vanadis system. As has been reported, there is additionally a cost-savings for medical systems using this technology over sequencing from a follow-up point of view due to the lower no call rate[10].

**Table three:** Comparison of Next-Generation Sequencing NIPT vs Vanadis^®^ NIPT

|  | **NGS NIPT<sup>5,15,22,34-36</sup>** | **Vanadis<sup>8,\*</sup>** |
| --- | --- | --- |
| **No call results** | 0.7 - 6.6% | 0.1-0.9% |
| **Sensitivity (21)** | 98.6 – >99.9% | >99.9% |
| **Sensitivity (18)** | 90 - >99.9% | 89 - >99.9% |
| **Sensitivity (13)** | 91.7 - >99.9% | >99.9% |
| **Specificity (21)** | 99.5 - 99.9% | 99.8 - >99.9% |
| **Specificity (18)** | 99.7 - >99.9% | 99.5% |
| **Specificity (13)** | 99.0 - 99.8% | 99.8 - >99.9% |

\*Including this study

Irrespective of the technology or methodology, there are some limitations to NIPT analysis which help to explain discrepancies between the test results and the fetal status. For example, since the cell-free fetal DNA is mainly produced by the placenta rather than fetus, false positive results can arise due to placental mosaicism[25-27] or the presence of a vanishing twin[28,29]. Additionally, false positive results or no call results may appear as a result of maternal cancer[30] or maternal chromosome anomalies[31]. Other limitations of the assay could arise from complex chromosomal abnormalities[26,32,33].

This study illustrates the high accuracy and clinical utility of Vanadis^®^ NIPT compared to traditional prenatal screening methods for common aneuploidy. As an equally accurate and reliable NIPT test, Vanadis^®^ NIPT can help eliminate the barrier to widespread usage of prenatal cfDNA for the global pregnancy population by being a technology that is significantly less complex to run and more cost effective.

## Supporting information

Validation Protocol

Validation Overview

## Data Availability

Data is available upon request

## ACKNOWLEDGEMENT

The authors would like to thank Professor Emmanuel Bujold and members of the Perinatal Biobank of the CHU de Québec-Université Laval for their assistance in collecting and preparing samples.

