## Supplementary material for "Assessment and clinical utility of a non-Next-Generation Sequencing based Non-Invasive Prenatal Testing technology": Validation Protocol

1. **PURPOSE**
   1.1 To provide a high degree of assurance that Vanadis NIPT System will produce accurate screening results for Trisomy 21, 18, and 13 and for fetal sex determination in the XXXXX laboratory.
2. **SCOPE**
   2.1 System Description
       2.1.1 Vanadis System - all equipment qualified for both installation qualification (IQ) and operational qualification (OQ).
       2.1.2 Vanadis Extract®
       2.1.3 Vanadis Core®
       2.1.4 Vanadis View ®
   2.2 The specimen type is blood collected in a 10 mL Streck® Cell-Free DNA BCT® and processed according to the manufacturer’s instructions to obtain the cfDNA. These instructions may be found in the Vanadis manual.
   2.3 Acceptable Sample Amount: 10 mL
   2.4 Preparation of the sample:
       2.4.1 Centrifuge at 1,300 x g for 30 minutes at 19 -25 °C.
       2.4.2 Transfer the plasma portion into a new centrifugation tube
             2.4.2.1 Do not disturb the buffy coat (lymphocyte layer)
       2.4.3 Centrifuge at 2,400 x g for 20 minutes at 19 -25 °C.
       2.4.4 Transfer the plasma fraction to plasma tubes (Sarstedt® Ref 62.611).
       2.4.5 Stability / Storage Conditions: 5 days, protect from light, 19 ‑ 25 °C (66 - 77 °F)
   2.5 Methodology
       2.5.1 The cell‑free DNA (cfDNA) used in this assay is extracted from the pregnant patient’s blood and contains not only the maternal cfDNA, but also a small fraction of placental cfDNA. The procedure consists of digestion of this cfDNA using restriction enzymes. The digested cfDNA is then hybridized and ligated to chromosome specific DNA probes forming a circular DNA. All non‑circular DNA is removed by exonuclease treatment. Finally, the circular DNA containing the cfDNA is amplified with rolling circle amplification to form rolling circle products (RCPs) that are labeled with chromosome‑specific fluorescently labeled DNA probes.
       2.5.2 The fluorescently labeled RCPs are deposited on the Vanadis View® Plate, image and counted with an automated microscopy scanner. The microscope takes multiple images from each well with different spectral filters, i.e., each wavelength range presents a specific chromosome. With image analysis algorithms, the fluorescently labeled RCPs are counted for each sample. The ratio between the number of chromosome‑specific RCPs is then transferred to risk calculation software, e.g., LifeCycle™ for Prenatal Screening to calculate the likelihood of a trisomy.
       2.5.3 Report Manager
       2.5.4 Data analysis and report generation
3. **DEFINITIONS**
   3.1 cfDNA	Cell free DNA
   3.2 NIPT	Non-invasive prenatal testing
   3.3 RCP	Rolling circle products
4. **ROLES AND RESPONSIBILITIES**

| Validation Plan Written by | XXXXX, Clinical Lab Manager |
| --- | --- |
| Validation Plan Review by | XXXXX, Head of Cytogenomics |
| Validation Plan Approval by | XXXXX, Director, Global Lab Operations |

5. **BACKGROUND / CLINICAL SIGNIFICANCE**
   5.1 Amplification - free targeted NIPT method to screen for trisomies 21, 18, 13 and to determine fetal sex in singleton pregnancies with automation.
6. **IMPACT ASSESSMENT**
   6.1 New methodology. All systems are impacted
       6.1.1 Safety
             6.1.1.1 See the attached Material Safety Data Sheets (MSDSs) for reagents.
       6.1.2 Equipment
       6.1.3 Reagents and Materials
             6.1.3.1 New supplies to order and establish inventory and ordering process
             6.1.3.2 Reagent qualification must be defined.
       6.1.4 Personnel
             6.1.4.1 Training and competency assessment needs developed.
       6.1.5 TRF
       6.1.6 Accessioning / LIMS
             6.1.6.1 Test codes:

Vanadis Common Aneuploidy Test (**XXXX**)

Vanadis Common Aneuploidy Test Plus Sex Determination (**XXXX**)

6.1.6.2 Train the accessioning staff.
6.1.7 Controlled documents
      6.1.7.1 SOPs needed for assay and equipment
      6.1.7.2 Equipment Preventive Maintainence (PM) logs
      6.1.7.3 Instrument manuals filed See attachments
6.1.8 Specimen Management
      6.1.8.1 Storage all frozen plasma stored at –70 °C to –80 °C (-94 to -112 °F).
      6.1.8.2 SeraCare reference materials for Trisomy 21, 18, and 13 stored at 2 ‑ 8 °C (36 - 46 °F).
6.1.9 Quality control
      6.1.9.1 Define quality control (QC) parameters and acceptance criteria, monitor
6.1.10 Proficiency Testing
       6.1.10.1 Establish proficiency testing (PT) or alternative laboratory proficiency assessment (ALPA) (TBD)
6.1.11 Reporting
       6.1.11.1 Design and approve new report
6.1.12 Quality indicators
       6.1.12.1 Turn around time (TAT)
6.2 Equipment, Reagents, and Materials
    6.2.1 Equipment / Materials

| **Materials and Supplies** | **Vendor** | **Manufacturer** | **Part/Cat #** | **Purpose** | **Storage Conditions** |
| --- | --- | --- | --- | --- | --- |
| Vanadis Extract® instrument | Vanadis | PerkinElmer® | 2026-0009 | To separate plasma and extract cfDNA | Temperature range during operation: 20‑24 °C (68 - 75 °F)  Operating relative humidity: 30‑60 % with no condensation |
| Vanadis Core® instrument | Vanadis | PerkinElmer® | 2027-0009 | Library prep and amplification for imaging | Temperature range during operation: 20‑24 °C (68 - 75 °F)  Operating relative humidity: 30 to 60 % with no condensation |
| Vanadis View® instrument | Vanadis | PerkinElmer® | 2028-0028 | To scan and analyze | Temperature range during operation: 20 ‑ 24 °C (68 -75 °F)  Operating relative humidity: 30 ‑ 60 % |
| Vanadis View® Plate | Vanadis | Wallac Oy | 4304-0010 | To capture labeled DNA molecules. | 19 - 25 °C (66 - 77 °F). Protect from dust and any other material to come into contact with the membrane. Handle with care. |
| Vanadis Core® Consumable kit | Vanadis | Wallac Oy | 4308-0010 | Consumable for Core® | 19 ‑ 25 °C (66 - 77 °F) |
| Vanadis Core® Reagent Cartridge Opening Tool | Vanadis | Wallac Oy | 61012813 | To open Reagent Cartridge | N/A |
| Vanadis Core® Reagent Cartridge Counter Balance | Vanadis | Wallac Oy | 4315-0010 | Balancer | N/A |
| Vanadis Extract® Sample Plates | Vanadis | Wallac Oy | 4305-0010 | Consumable for Vanadis Extract® | 2 ‑ 30 °C (36 - 86 °F) |
| Vanadis Extract® Reaction Plates | Vanadis | Wallac Oy | 4306-0010 | Consumable for Vanadis Extract® | 2 ‑ 30 °C (36 - 86 °F) |
| Vanadis Extract® Sleeves and Tubes | Vanadis | Wallac Oy | 4307-0010 | Consumable for Vanadis Extract® | N/A |
| Vanadis Extract® Weekly Maintenance Tool | Vanadis | Wallac Oy | 2026-4040 | Vanadis Extract® maintenance use | N/A |
| Tubes for Vanadis Extract® weekly maintenance | Vanadis | Wallac Oy | 2026-4050 | Vanadis Extract® maintenance use | N/A |
| Sarstedt® 5 mL plasma storage tube | Unimed Healthcare SB | Sarstedt, Inc | 62.611 | Plasma storage | N/A |
| 4 mL disposable tips with filter in tip rack | Vanadis | Wallac Oy | 2026-4020 | Consumable for Vanadis Extract® | –15 to 55 °C (5 - 131 °F) |
| 1,000 μL disposable tips with filter in tip rack | Vanadis | Wallac Oy | 2027-4010 | Consumable for Vanadis Extract® and Core® | –15 to 55 °C (5 - 131 °F) |
| 10 ‑ 300 μL disposable tips with filter in tip rack | Vanadis | Wallac Oy | 2027-4020 | Consumable for Vanadis Core® | –15 to 55 °C (5 - 131 °F) |
| 0.5 ‑ 50 μL disposable tips without filter in nested tip rack | Vanadis | Wallac Oy | 2027-4030 | Consumable for Vanadis Core® | –15 ‑ 55 °C (5 - 131 °F) |

6.2.2 Reagents

| **Reagent** | **Vendor** | **Manufacturer** | **Part/Cat#** | **Purpose** | **Storage Conditions** |
| --- | --- | --- | --- | --- | --- |
| Vanadis Core®Chromosome  Quantification kit  Consists of 2 packages  Reagent Cartridge  Exp: As stated on the outer label  Store frozen and protected from light in original package  Before use transfer to 2 ‑ 8 °C (36 - 46 °F)  Exp: 3 days, do not refreeze  Restriction enzyme (57.5 µL)  Oligo mix - mixture of oligonucleotides in TRIS-EDTA buffer (540 µL)  DNA ligase enzyme (22.5 µL)  NAD (Nicotinamide adenine dinucleotide) solution (235 μL)  Exonuclease enzyme  1 (390 µL), 2 (125 µL),  3 (60 µL) and 4 (57.5 µL)  Water (1,225 µL)  RCA primer - oligonucleotides in salt buffer (1,040 µL)  dNTP Solution (mixture of deoxynucleotides in salt butter (360 µL)  DNA polymerase enzyme (112.5 µL)  Detection Oligos Fluorescently labeled oligonucleotides in salt buffer (320 µL) | Vanadis | Wallac Oy | 4302-001B | Assay | –30 to 16 °C (-22 to 61 °F) |
| Buffer 1 (5 x 300 µL), 2 (3 x 300 µL)  DNA control material (A, B, C)  (3 x 210 µL) | Vanadis | Wallac Oy | 4302-001B | Assay | –30 to 16 °C (-22 to 61 °F) |
| Labeling Buffer  (2 tubes, 1.1 mL)  Exp: As stated on the outer label | Vanadis | Wallac Oy | 4302-001B | Assay | 19 ‑ 25 °C (66 - 77 °F) |
| Vanadis Extract® Reagent kit  Consists of two packages  P1  Proteinase K  (8 tubes, 1.0 mL)  Magnetic Beads  (8 tubes, 0.4 mL)  Elution Buffer  (16 tubes, 1.9 mL)  Exp: As stated on the outer label | Vanadis | Wallac Oy | 4300-0010 | Extraction of cell-free DNA from plasma | P1: 2 ‑ 8 °C (36 - 46 °F) |
| P2  SDS Solution  (16 tubes, 1.9 mL)  Exp: As stated on the outer label |  |  |  |  | P2: 19 ‑ 25 °C (66 - 77 °F) |
| Vanadis Extract® Bind and Wash Kit  Consists of three packages  P1  Binding Buffer  (1 bottle, 640 mL) x 2  P2  Wash Solution  (2 bottles, 750 mL)  Exp: As stated on the outer label | Vanadis | Wallac Oy | 4301-0010 | For further purification of extracted cell-free DNA | 19 - 25 °C (66 - 77 °F) |
| Vanadis Core® Buffer Kit  Wash Buffer  (2 troughs, 60 mL)  Prep: Ready to use  Exp: As stated on the trough label  Diluent Buffer  (1 trough, 45 mL)  Prep: Ready to use  Exp: As stated on the trough label  Detergent (Tween® 20)  (1 trough, 50 mL)  Prep: Ready to use  Exp: As stated on the trough label | Vanadis | Wallac Oy | 4303-0010 | Diluting exonuclease reactions and for diluting and removing excel reagents from the labeled DNA molecular mix | 19 ‑ 25 °C (66 - 77 °F) |

7. **TEST DESCRIPTION AND TEST PLAN**
   7.1 Test plan: Recommendation of 5 runs (with a minimum of 3) after training and competency established by the vendor. As part of competency runs, 48 pooled plasma samples were run on the Vanadis Extract®, Core®, and View®.
       7.1.1 Staff competency run
       7.1.2 Total of 48 samples
   7.2 Validation Runs: planned with convenient sample amount for 98% confidence level and 95% reliability.  At least three runs should be performed. Samples from known affected pregnancies can be used in place of SeraSeq samples.
       7.2.1 Validation Run A
             7.2.1.1 Total of 84 samples
                     7.2.1.1.1 2 SeraSeq T18
                     7.2.1.1.2 1 SeraSeq T13
                     7.2.1.1.3 1 SeraSeq T21
                     7.2.1.1.4 80 normal plasma samples (pregnancy)
                     7.2.1.1.5 Ideally, a random mix of plasma samples from pregnant women should be run. For samples preselected for a designed run, a sex ratio of 60:40 is tolerated
       7.2.2 Validation Run B
             7.2.2.1 Total of 84 samples
                     7.2.2.1.1 2 SeraSeq T21
                     7.2.2.1.2 1 SeraSeq T13
                     7.2.2.1.3 1 SeraSeq T18
                     7.2.2.1.4 80 normal plasma samples (pregnancy), 34 females and 46 males
       7.2.3 Validation Run C
             7.2.3.1 Total of 84 samples
                     7.2.3.1.1 1 SeraSeq T21
                     7.2.3.1.2 2 SeraSeq T13
                     7.2.3.1.3 1 SeraSeq T18
                     7.2.3.1.4 80 normal plasma samples (pregnancy), 34 female fetuses and 46 male fetuses
       7.2.4 Validation Run D
             7.2.4.1 Total of 84 samples
                     7.2.4.1.1 1 SeraSeq T18
                     7.2.4.1.2 1 SeraSeq T13
                     7.2.4.1.3 2 SeraSeq T21
                     7.2.4.1.4 80 normal plasma samples (pregnancy), 34 female fetuses and 46 male fetuses
       7.2.5 Validation Run E
             7.2.5.1 Total of 72 samples
                     7.2.5.1.1 2 SeraSeq T21
                     7.2.5.1.2 1 SeraSeq T18
                     7.2.5.1.3 1 SeraSeq T13
                     7.2.5.1.4 68 normal plasma samples (pregnancy), 28 female fetuses, 40 male fetuses.
       7.2.6 Validation Run F
             7.2.6.1 Total of 72 samples
                     7.2.6.1.1 2 SeraSeq T21
                     7.2.6.1.2 1 SeraSeq T18
                     7.2.6.1.3 1 SeraSeq T13
                     7.2.6.1.4 68 normal plasma samples (pregnancy), 28 female fetuses, 40 male fetuses.
   7.3 Quality control
       7.3.1 Plate quality metrics must pass for each run.
       7.3.2 QC monitored
             7.3.2.1 Batch QC must pass
             7.3.2.2 Median density of the samples within each plate must be within 7.78 ‑ 9.84.
             7.3.2.3 The Coefficient of Variation (CV) of the ratio for each batch is within acceptable limits and batch to batch is monitored.
       7.3.3 Plate ratio
             7.3.3.1 Sample mean ratio 0.98 ‑ 1.02
             7.3.3.2 Density Median Log counts per image for chromosomes 13, 18 and 21: 7.77 to 9.84
       7.3.4 PERFORMANCE CRITERIA AND EXPECTED RESULTS
             7.3.4.1 Validation/Verification Plan with Expected Results
             7.3.4.2 For Aneuploidies

| **PERFORMANCE CRITERIA** | **DEFINITION** | **EXPECTED RESULTS** |
| --- | --- | --- |
| ACCURACY for Chromosomes 21, 13, 18 independently | (TP + TN)/All Results | ≥ 0.90 |
| PRECISION for chromosomes 21, 18, 13 independently | TP/TP + FP | ≥ 0.90 |
| ANALYTICAL SPECIFICITY for Chromosomes 21, 13, 18 independently | TN/TN + FP | ≥ 0.90 |
| ANALYTICAL SENSITIVITY for Chromosomes 21, 13, 18 independently | TP/TP + FN | ≥ 0.90 |
| Positive Predictive Value (Precision) for chromosomes 21, 18, 13 combined | TP/TP + FP | ≥ 0.90 |
| Negative Predictive Value (NPV) for Chromosomes 21, 13, 18 independently | TN/TN + FN | ≥ 0.90 |
| False Negative Rate (FNR) for Chromosomes 21, 13, 18 independently | FN/FN + TP | < 0.10 |
| False Positive Rate (FPR) for Chromosomes 21, 13, 18 independently | FP/FP + TN | < 0.10 |

7.3.4.3 For Sex Determination (gender calls)

| **PERFORMANCE CRITERIA** | **DEFINITION** | **EXPECTED RESULTS** |
| --- | --- | --- |
| ACCURACY | (TP + TN)/All Results | ≥ 0.90 |
| PRECISION | TP/TP + FP | ≥ 0.90 |
| ANALYTICAL SPECIFICITY | TN/TN + FP | ≥ 0.90 |
| ANALYTICAL SENSITIVITY | TP/TP + FN | ≥ 0.90 |

7.3.5 Failure Criteria:
      7.3.5.1 instrument failures that lead to loss of date would be qualified as a "complete fail run".
      7.3.5.2 If greater than 5% of samples in a run have no results, please contact PerkinElmer to discuss how to proceed.
      7.3.5.3 If plate QC metrics fail or plate controls fail- the run will be “failed”.
7.4 REFERENCES AND LINKS
    7.4.1 Vanadis Core® T21/T18/T13 Reagent Cartridge, November 2018
    7.4.2 Vanadis Core® Chromosome Quantification Kit, December 2018
    7.4.3 Vanadis Extract® Reagent Kit, January 2019
    7.4.4 Vanadis Extract® Bind and Wash Kit, December 2018
    7.4.5 Vanadis Core® Buffer Kit, December 2018
    7.4.6 Vanadis View® Plate, September 2018
    7.4.7 Vanadis Core® Instrument Manual, 2026-9030-01 January 2019
    7.4.8 Vanadis Extract® Instrument Manual, 2026-9030-01 January 2019
    7.4.9 Vanadis Molecular Counting Unit® Instrument Manual, 2028-9050-01 January 2019

8. **DOCUMENT CONTROL**

| **Role** | **Personnel** |
| --- | --- |
| Owner |  |
| Writers |  |
| Reviewers |  |
| Approvers |  |

9. **DOCUMENT HISTORY**

| Version | **Summary of Changes** | **Date** |
| --- | --- | --- |
| 1 | New Document. |  |
