## Supplementary material for "Assessment and clinical utility of a non-Next-Generation Sequencing based Non-Invasive Prenatal Testing technology": Validation Overview

**Vanadis NIPT Assay Validation Plan**

**Background:**

Non-invasive prenatal testing (NIPT) is based on the analysis of placental cell-free DNA (cfDNA) circulating in maternal blood and provides a highly accurate screening option for trisomy 21 (T21, Down syndrome), trisomy 18 (T18, Edwards syndrome), trisomy 13 (T13, Patau syndrome) and fetal sex, thus reducing the need for invasive testing.

The automated Vanadis NIPT workflow consists of three main stages: **Vanadis Extract®** for cfDNA extraction, **Vanadis Core®** for the formation of rolling circle replication products, and **Vanadis View®** for the molecule counting**.** All three stages are controlled through the Vanadis System Software**®** which guides the workflow and monitors the assay performance by conducting the built-in Vanadis® NIPT quality assessment procedures. After quality assessment, the results are sent to a second software for trisomy risk calculation and reporting i.e. **LifeCycle™**.

Vanadis technology converts target chromosomes into digitally quantifiable objects. It is the only screening assay targeting specific chromosomes without using PCR amplification. Instead, target fragments are directly captured using probes before being converted to circular DNA objects that are replicated to form DNA bundles and are labeled for counting. Eliminating PCR improves the assay precision and removes the requirement to separate the workflow into pre- and post-PCR areas. In addition, by targeting thousands of chromosomal positions, the Vanadis platform quantifies, on average, 650,000 molecules per chromosome. This is more than three-fold greater than sequencing technology. The high precision achieved through incorporating high-yield counting and elimination of PCR improves the detection rate and reduces the no-call rate using the Vanadis method.

**Purpose:**

The Vanadis NIPT assay (PerkinElmer) validation at the laboratory is planned to compare the results of this method against samples with known outcomes for Trisomies 13, 18 and 21. Both screening methods use cfDNA extracted from blood specimens drawn from pregnant patients.

**Performance metric evaluation:** The goal of the validation study is to evaluate the performance metrics of the Vanadis assay. The following metrics will be evaluated based on the Z-score results from Vanadis. The manufacturer’s Z-score cut-offs and thresholds will be used for the post-test risk calculation and interpretation of the Vanadis NIPT test.

1. Sensitivity
2. Specificity
3. Accuracy
4. Precision
5. False positive rate
6. False negative rate

**Specimen collection process for validation study**

In order to collect specimens for the validation study, several physician offices in this network were approached and asked to participate in the specimen consent and collection process. In order to participate, each office was required to identify a study coordinator. Study coordinators were required to be certified in protecting human research participants; those who did not currently hold this certification were required to successfully complete a training course and obtain their certification.

A draft consent form and study protocol was developed and presented to an Institutional Review Board (IRB) for approval. IRB approval was secured and the regulatory-required study binders were then provided to each office/study coordinator. Eligible patients (those who were planning to undergo NIPT testing already through the laboratory) were approached to ask if they would be willing to participate in the study. A copy of the consent form was provided; the study coordinator was available to answer any questions the patients may have about the study. Patients who agreed to participate were required to sign the consent form, which was also signed by the study coordinator. A copy of the consent form was provided to the patient, with a copy saved in the study binder. The physicians then added a note to the patient’s NIPT lab order, only for patients who provided their consent, requesting that additional specimens (2 Streck tubes of blood - 10 ml each) be collected at the time the patient was having her blood drawn for her regular NIPT. The additional specimens collected for validation were marked accordingly and were sent to the lab for use in the validation process. The primary specimens collected were sent to the outside laboratory for the patients’ standard NIPT to be performed.

Summary:

- Total number of patient specimens planned for collection and validation: N=240.
- 4 tubes of blood collected per patient, with 2 sent to the outside laboratory for their standard NIPT testing and 2 sent to our laboratory for Vanadis testing.

**PROCEDURE:**

Under the IRB umbrella, 2 extra tubes of blood will be collected from pregnant patients from various associated clinics in the region for the Vanadis NIPT validation study. Plasma will be extracted from both tubes using Vanadis extract protocols and frozen until ready for further processing. The blood collected from the second tube will be used as a backup for any test failures and repeats during the validation process.  The study will commence when the first set of 80 frozen plasma specimens are extracted and ready for testing. The total number of patient specimens planned for collection is at least 240.

Validation study will strictly adhere to the following standard operating procedures (SOPs) that have been approved and signed by personnel. Trained medical technologists will be assigned to carry out the validation runs using qualified reagents and certified instruments.

- Vanadis NIPT Extract SOP
- Vanadis NIPT Core SOP
- Vanadis NIPT View image and QC Analysis SOP
- Vanadis NIPT Risk Analysis and Reporting SOP

Each run will use at least 4 positive controls, including at least one each of Trisomy 21, 18 and 13. If previously confirmed positive controls are not available, these controls will be purchased from Seracare.

<https://www.seracare.com/Seraseq-Trisomy-21-Aneuploidy-Reference-Material-0720-0019/>

<https://www.seracare.com/Seraseq-Trisomy-18-Aneuploidy-Reference-Material-0720-0018/>

<https://www.seracare.com/Seraseq-Trisomy-13-Aneuploidy-Reference-Material-0720-0017/>

Ideally, a random mix of plasma samples from pregnant women should be run. For samples preselected for a designed run, a sex ratio of 60:40 is tolerated.

**Validation will be carried out in 2 parts as outlined below:**

1. **Blind study:**

Total of at least 240 blood samples drawn under the IRB umbrella will be processed for the Blind study. This study will address performance metrics as defined in the “Purpose” section of this document. Study will commence when the first set of 80 frozen plasma specimens are separated and ready for testing.

The blind study will consist of 3-5 runs of specimens from pregnant patients, split and run in parallel with the outside for their routine NIPT testing. Samples will be run on the Vanadis assay and compared with the previously validated results from the standard screening method (outside NIPT).

- Maternal blood collected
  - 4 Streck blood tubes of ~10ml each, from pregnant patients
    - Two tubes to be sent to the outside laboratory for their routine NIPT testing.
    - 3rd tube will be run on the Vanadis assay and compared with the outside laboratory’s screening method results.
    - 4th tube will be kept for any repeats and reproducibility/ inter-run comparisons
- No less than 3 runs of 80 patient specimens each
- Total 84 samples per run
  - 80 patient specimens
  - 4 reference material/controls purchased from Seracare (at least one each of 13, 18 and 21 positive)

NOTE: If the 3 runs as described above are highly successful with zero “No calls” and no discrepant results against the standard NIPT assay that need resolution, then the blind study will be deemed complete with 240 total samples. Any remaining specimens will be used for an extra run if necessary, as determined by the Lab Director. If more than 5% of the results are discrepant, PerkinElmer will be contacted to discuss how to proceed.

**Acceptance Criteria:**

- Vanadis QC metrics/controls: Quality metrics defined in SOP for “Vanadis View Image and QC analysis” for Kit Controls, Patient Samples and the Assay Run are met as per specifications
  - Run CV’s will be monitored for each validation run to assess data quality and interpretation of results.
  - Performance of the Vanadis kit controls will be monitored for each validation run
- Final outcomes (Negative, Positive and Inconclusive) from all samples should be 95% concordant with external laboratory. Any discrepancies observed should be resolved and documented.

**Discordant results resolution:**

Discordant results between an alternate technology platform and Vanadis NIPT platforms:

- Different NIPT methods have different limitations and sensitivity and need to be considered in case of discordant results
- In case of discordant results between 2 NIPT assays, appropriate investigation needs to be performed to document the results
- Sensitivity and specificity calculations will be adjusted accordingly to resolve the discrepancy
- Performance metrics will be assessed by gold standard methods (1) Invasive diagnostic testing by karyotyping/FISH/microarray (Amnio/CVS) or (2) Pregnancy/birth outcome

**Interpretation considerations:**

- Placental mosaicism needs to be considered when interpreting/comparing NIPT results
- Other biological factors such as vanishing twins, etc. need to be considered when interpreting/comparing NIPT results

2. **Reproducibility Study:**

To further validate the Vanadis NIPT assay, inter runs can be performed to demonstrate data reproducibility.

The reproducibility study would consist of 1 run using the second tube that was not used for repeat runs during the blind study. This study would be limited to at least 45 samples for inter-assay reproducibility.

Total of at least 48 samples =45 blood draws plus 3 controls purchased from Seracare (one positive each for Trisomy 13, 18 and 21)

**Limitation**: Due to limited availability of cfDNA for a given specimen, **Intra**-assay reproducibility is not a viable option for this method.

**Acceptance Criteria:**

- Vanadis QC metrics/controls: Quality metrics defined in SOP for “Vanadis View Image and QC analysis” for Kit Controls, Patient Samples and the Assay Run are met as per specifications
  - Run CV’s will be monitored for each validation run to assess data quality and interpretation of results.
  - Performance of the Vanadis kit controls will be monitored for each validation run
- Final outcomes (Negative, Positive and Inconclusive) based on Z-score calculations for all specimens should match those from the blind study runs for the same specimens that have been replicated in this study

**Signatures and approvals**

Medical Technologist	_________________________	Date: _____________

Laboratory Supervisor _________________________	Date: _____________

Approved By:	_________________________	Date: _____________
